# An open-access database of infectious disease transmission trees to explore superspreader epidemiology

**DOI:** 10.1101/2021.01.11.21249622

**Authors:** Juliana C. Taube, Paige B. Miller, John M. Drake

## Abstract

Historically, emerging and re-emerging infectious diseases have caused large, deadly, and expensive multi-national outbreaks. Often outbreak investigations aim to identify who infected whom by reconstructing the outbreak transmission tree, which visualizes transmission between individuals as a network with nodes representing individuals and branches representing transmission from person to person. We compiled a database of 383 published, standardized transmission trees consisting of 16 directly-transmitted diseases ranging in size from 2 to 286 cases. For each tree and disease we calculated several key statistics, such as outbreak size, average number of secondary infections, the dispersion parameter, and the number of superspreaders. We demonstrated the potential utility of the database through short analyses addressing questions about superspreader epidemiology for a variety of diseases, including COVID-19. First, we compared the frequency and contribution of superspreaders to onward transmission across diseases. COVID-19 outbreaks had significantly fewer superspreaders than outbreaks of SARS and MERS and a dispersion parameter between that of SARS and MERS. Across diseases the presence of more superspreaders was associated with greater outbreak size. Second, we further examined how early spread impacts tree size. Generally, trees sparked by a superspreader had larger outbreak sizes than those trees not sparked by a superspreader, and this trend was significant for COVID-19 trees. Third, we investigated patterns in how superspreaders are infected. Across trees with more than one superspreader, we found support for the theory that superspreaders generate other superspreaders, even when controlling for number of secondary infections. In sum, our findings put the role of superspreading to COVID-19 transmission in perspective with that of SARS and MERS and suggest an avenue for further research on the generation of superspreaders. These data have been made openly available to encourage reuse and further scientific inquiry.

**Author Summary:** Public health investigations often aim to identify who infected whom, or the transmission tree, during outbreaks of infectious diseases. These investigations tend to be resource intensive but valuable as they contain epidemiological information, including the average number of infections caused by each individual and the variation in this number. To date, there remains no standardized format nor comprehensive database of infectious disease transmission trees. To fill this gap, we standardized and compiled more than 350 published transmission trees for 16 directly-transmitted diseases into a database that is publicly available. In this paper, we give an overview of the database construction process, as well as a demonstration of the types of questions that the database can be used to answer related to superspreader epidemiology. For example, we show that COVID-19 outbreaks have fewer superspreaders than outbreaks of SARS and MERS. We also find support for the theory that superspreaders generate other superspreaders. In the future, this database can be used to answer other outstanding questions in the field of epidemiology.

## Introduction

In the past 20 years, emerging and re-emerging infectious diseases have caused large, deadly, and expensive multi-national outbreaks of SARS-CoV (SARS), Zika, Ebola, measles and now SARS-CoV-2 (COVID-19). From these outbreaks, researchers have learned a great deal about the epidemiology of each disease. For example, spread of SARS in 2003 was greatly facilitated by superspreaders, individuals that infect an unusually high number of contacts, especially on airplanes and in health care settings [1,2]. During the present COVID-19 pandemic, numerous instances of superspreading events in churches, choir practices, exercise classes, weddings, and other settings have been reported [3]. Understanding the factors leading to excess transmission by some individuals or in some settings is a major goal of outbreak investigations and may lead to improved control strategies.

Some outbreak investigations aim to identify *who infected whom* by reconstructing the outbreak *transmission tree*, which visualizes transmission between individuals as networks with nodes representing individuals and branches representing transmission from person to person. Transmission trees are typically investigated by case-finding, contact-tracing, and detailed epidemiological interviews followed sometimes by probabilistic reconstruction [4]. These investigations are costly but necessary because transmission trees contain information about key epidemiological parameters including the outbreak size, the average number of secondary infections (or *reproduction number, R*), and the dispersion parameter (variation in *R*). These parameters govern the effectiveness of disease control strategies and can be used to inform models of spread [5, 6]. Additionally, comparing the statistics of different transmission trees can lead to insights about how transmission differs across the contexts of pathogen, time, and place. We compiled and standardized transmission trees from a wide variety of sources for general use by the research community. Using the database, we test several hypotheses addressing aspects of superspreader epidemiology.

In 2005, Lloyd-Smith and colleagues [6] proposed a definition for superspreading based on variation in the offspring distribution (e.g. the number of infections caused by each infected individual). According to this proposal, superspreaders are cases causing more secondary infections than the 99th percentile of a Poisson(*R*_0_) distribution. In their analysis, superspreading was especially important for understanding the offspring distribution of SARS and measles. With these findings in mind, we used the database to examine three hypotheses about the frequency of superspreading in COVID-19 outbreaks relative to other diseases.

1. We hypothesized that there would be a positive relationship between number of superspreaders and tree size.
2. We hypothesized that COVID-19, as a coronavirus, would have a similar dispersion parameter to that of SARS and MERS, but lower than that of other diseases not typically associated with superspreading, such as influenza.
3. We hypothesized that superspreaders would have directly and indirectly caused a larger cumulative proportion of cases in outbreaks of COVID-19, SARS, and MERS than diseases where superspreading is less common, such as influenza and measles. Lloyd-Smith and colleagues [6] also posited that diseases that have larger variation in offspring distributions (i.e. lower dispersion parameters) have a greater chance of extinction which increases further as *R*_0_ declines toward 1. However, early superspreading events may prevent extinction by increasing the size from which the outbreak grows and making infection propagation more likely [7]. Additionally, the probability of *detecting* an outbreak may be higher if there is a superspreading event, because public health officials are more likely to investigate a cluster than an isolated case. For these reasons, in our second analysis we test two hypotheses relating tree size and early spread.
4. We hypothesized that trees sparked by superspreaders would be larger than trees not sparked by superspreaders.
5. We hypothesized that the proportion of cases in the first generation would be positively associated with standardized tree size. While it is clear that superspreading helps explain the propagation patterns of several infectious diseases [6], how superspreaders themselves are generated (i.e., who spreads to superspreaders) is poorly understood. A new hypothesis for COVID-19 suggests that superspreaders may generate new superspreaders via a mechanism based on initial viral dosage [8] if superspreaders are individuals with unusually high viral shedding because they themselves were exposed to other individuals with high viral shedding. Another explanation for superspreaders generating new superspreaders could be that superspreaders engage in riskier behavior and are more likely to infect others with similar behavior. For example, superspreaders and their contacts may be more likely to attend large gatherings or less likely to take precautionary measures to mitigate spread (e.g. masks) than the general public. These proposed mechanisms suggest that generation of superspreaders is not random, but correlated with characteristics of the superspreader’s infector or tied to the context of transmission. We investigate these patterns of superspreader generation in our third analysis.
6. We hypothesized that superspreaders would be more likely to infect other superspreaders, controlling for number of secondary infections.

Here, we document the construction of OutbreakTrees and demonstrate the potential utility of this database by investigating three features of superspreading epidemiology: frequency, timing, and generation.

## Results & Discussion

### Database summary statistics

Currently, OutbreakTrees includes 383 trees describing 16 directly-transmitted infectious diseases (see Fig 1 for examples), most of which are caused by viruses (Fig 2). COVID-19 outbreaks comprise 256, or approximately 67%, of the trees in the database. Trees range in size from 2 to 286 individuals. This database contains data for outbreaks that took place in the years 1946 through 2020. The most common node attributes for trees include context of transmission (work, school, family, etc.), date of onset, sex, and age (Table 1).

**Table 1:** List of most common attributes for individuals in trees.

| Attribute | Database Code | Number of Trees |
| --- | --- | --- |
| Transmission context | cont | 301 |
| Symptom Onset | onset | 137 |
| Sex | sex | 86 |
| Age | age | 69 |
| Location | loc | 56 |
| Quarantine Status | quar | 36 |
| Occupation | occp | 34 |
| Survival | surv | 20 |

**Figure 1:**
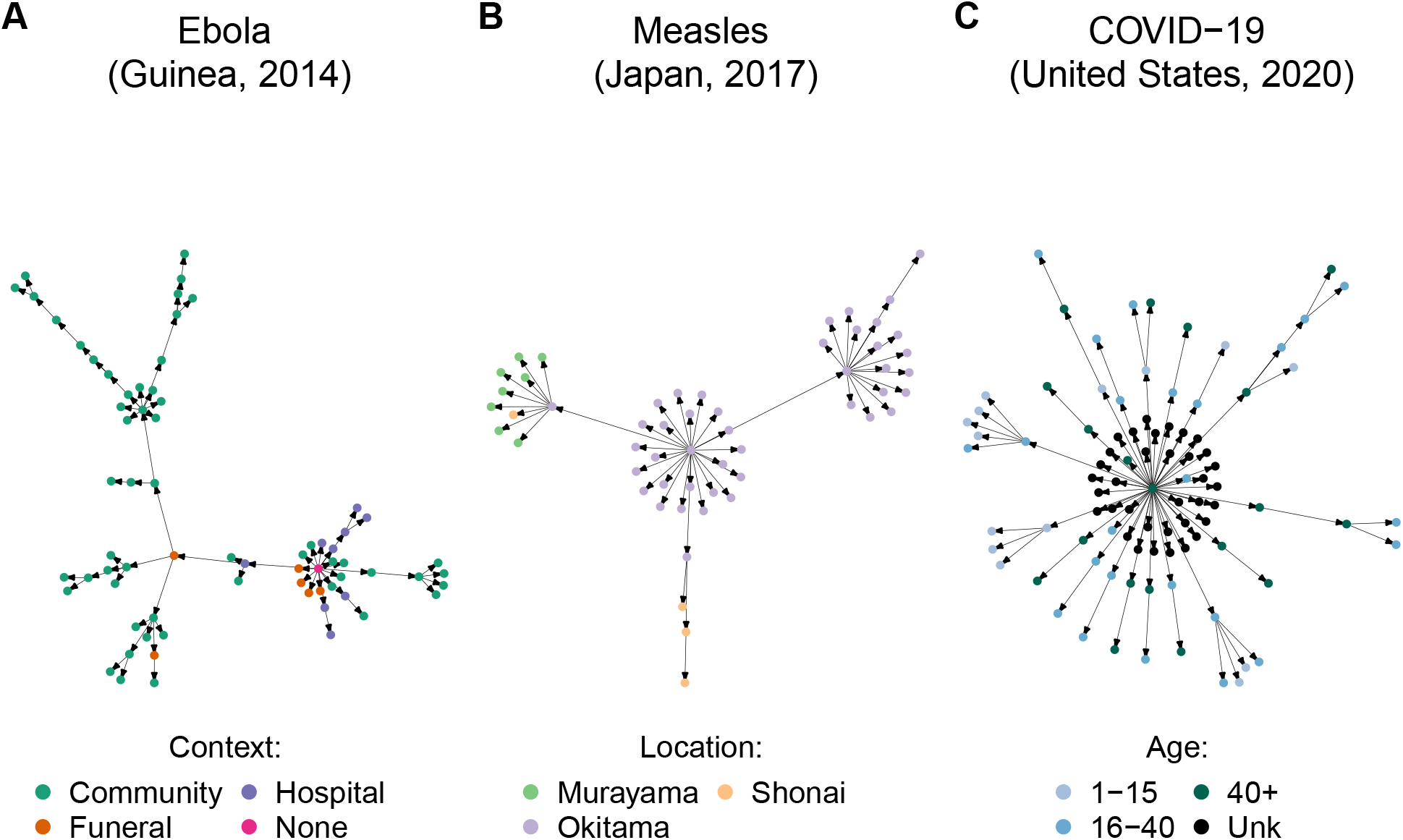
We compiled infectious disease transmission trees from the literature along with reported attribute information. Shown here are example trees in the database. (A) Ebola spread in different contexts [9]. (B) Measles spread in different locations [10]. (C) COVID-19 spread among age classes [11]. Primary sources for transmission trees are available in OutbreakTrees and listed in the Supplemental Material. OutbreakTrees may be accessed online at OutbreakTrees.ecology.uga.edu

**Figure 2:**
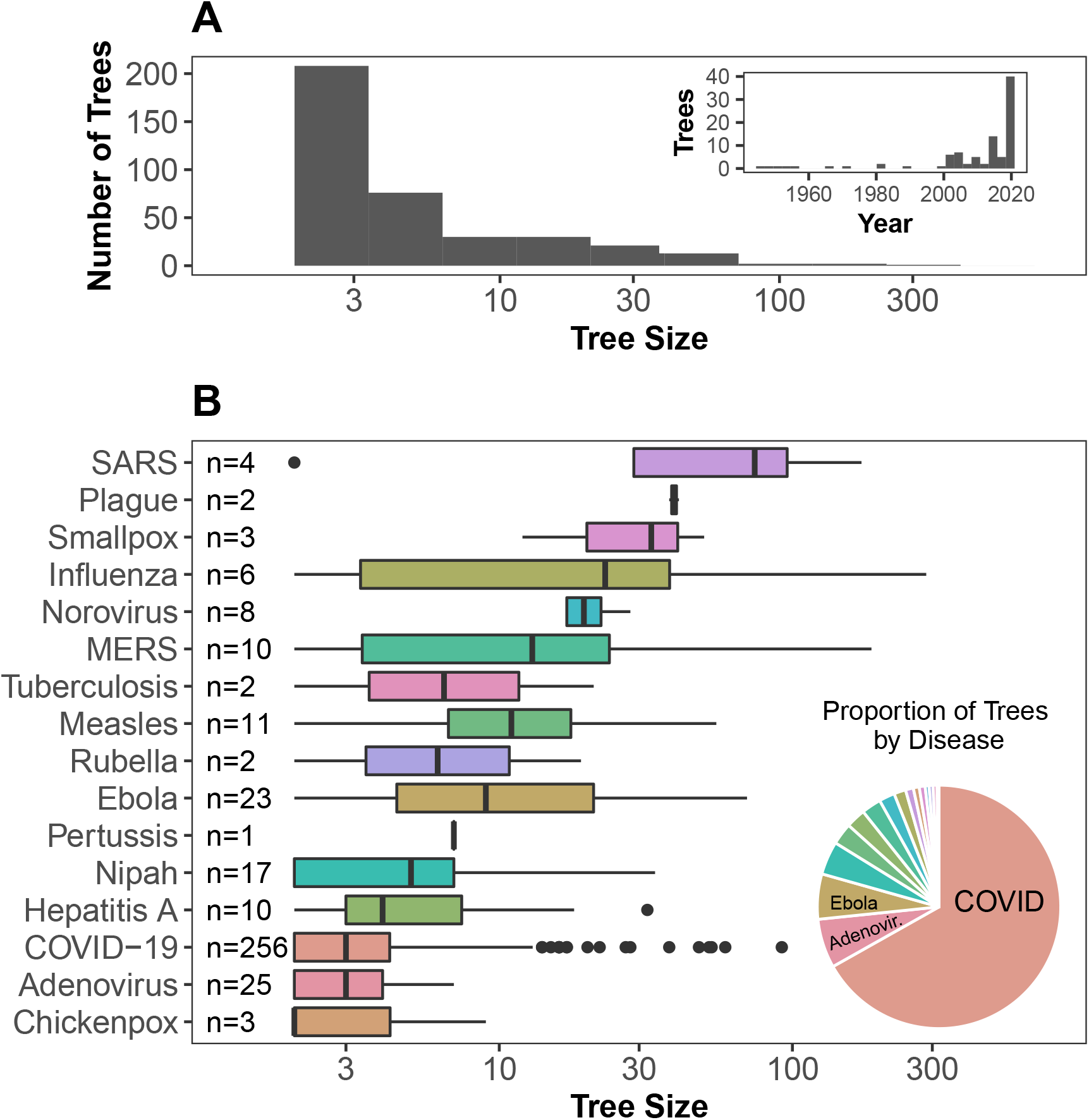
Characteristics of transmission trees in OutbreakTrees. (A) Tree size varies from 2 to 286 with a median of 3 and most trees represent outbreaks taking place in the past 20 years. (B) The largest trees are from H1N1 and SARS outbreaks while the highest proportion of trees in the database are from outbreaks of COVID-19, followed by adenovirus and Ebola. Tree size axes in both plots are shown on a log10 scale to better illustrate variation in medium-sized trees.

### Superspreading events across diseases (Hypotheses 1-3)

Consistent with our first hypothesis, larger trees were associated with more superspreaders (Fig 3A). Though outbreaks of COVID-19 contained fewer superspreaders than outbreaks of SARS (P=0.00043) and MERS (P=0.000036) (Wilcoxon test with Bonferroni-Holm correction, S3 Fig), COVID-19 had a median dispersion parameter (*k* = 0.1) (Fig 3B) between that of SARS and MERS, supporting our second hypothesis, and in line with some previous estimates ([12] but see [13, 14, 15, 16, 17]) which give a range of estimates across these coronaviruses). Lastly, addressing our third hypothesis, superspreaders were found to directly or indirectly account for all cases in at least 50% of tuberculosis, SARS, rubella, MERS, Ebola, and COVID-19 outbreaks (Fig 3C). The lowest median percentages of 0 were observed in chickenpox, hepatitis A, and norovirus trees.

**Figure 3:**
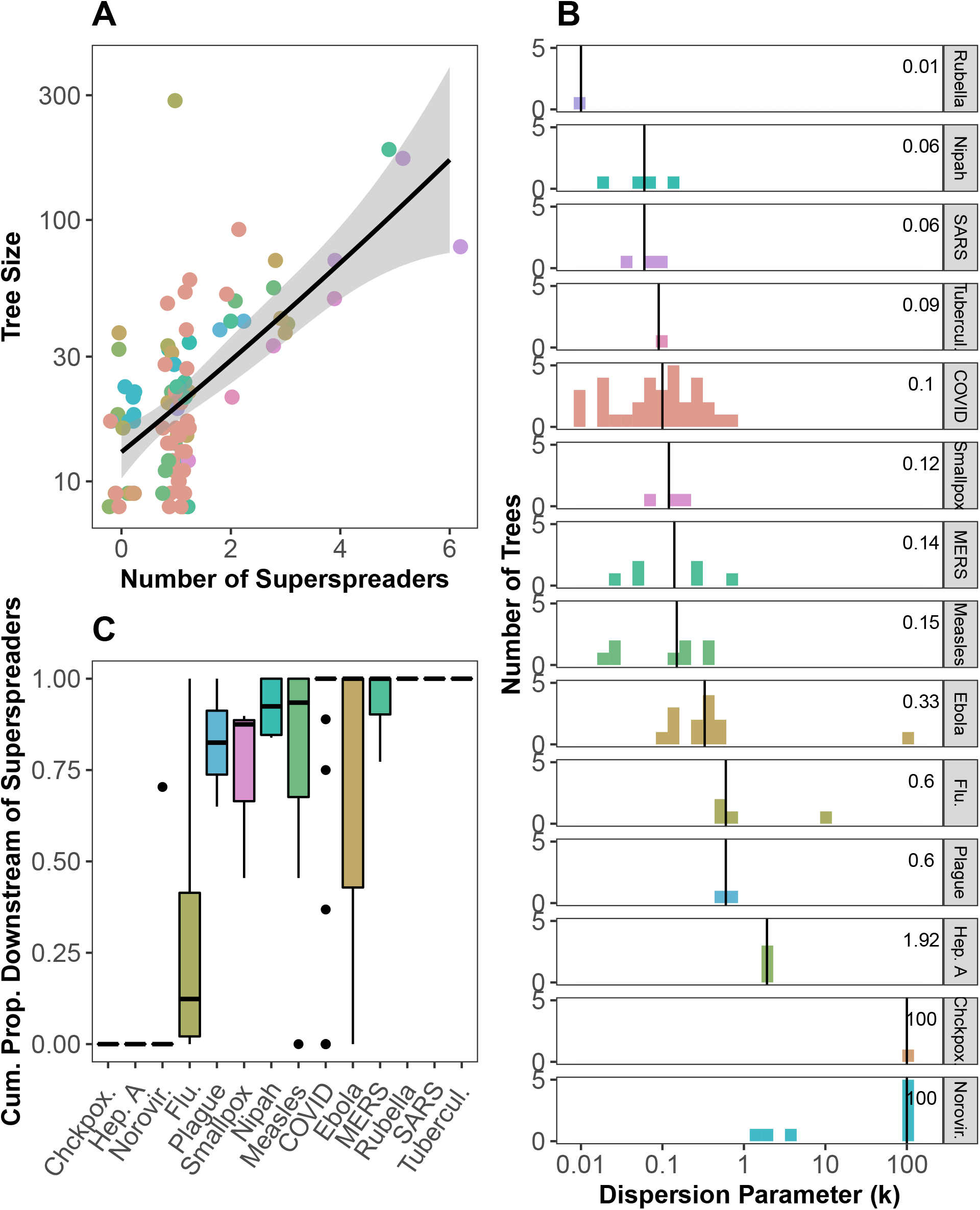
The importance of superspreading across diseases measured in three ways. (A) Larger outbreaks tend to have more superspreaders. Points are jittered vertically and y-axis is on a log10 scale for visual aid. (B) Dispersion parameter (*k*) of a negative binomial distribution fit to the offspring distribution of trees by disease. Vertical line and value printed in each facet shows the median *k* for each disease. (C) Cumulative proportion of cases directly or indirectly attributed to superspreaders in a tree by disease. Only trees with 8 or more cases were used in these analyses with other cutoffs shown in S1 Fig and S2 Fig.

Taken together, these results indicate that COVID-19 is able to reach large numbers of individuals with relatively few superspreaders (compared with SARS and MERS) that account for a large proportion of onward transmission. These results, combined with a dispersion parameter in between that of SARS and MERS, put the role of superspreading in COVID-19 trees into perspective: superspreaders contribute substantially to onward transmission of COVID-19 but in a way that seems distinct from SARS and MERS. In particular, the high downstream effects of COVID-19 cases caused by relatively few superspreaders could reflect important sampling differences in COVID-19 trees such as superspreading disproportionately increasing detection relative to other diseases. We investigate patterns associated with early spread in the next section.

### Tree size and early spreading events (Hypotheses 4-5)

In our database, the proportion of trees sparked by superspreaders varied by disease; half of MERS trees (5/10) and nearly half of Ebola (11/23) and Nipah (7/17) trees were sparked by superspreaders. Less than one-third of measles (3/11), COVID-19 (40/256), influenza (1/5), and adenovirus (1/25) trees were initiated by superspreaders. None of the 8 norovirus trees or 10 hepatitis A trees were sparked by superspreaders (Fig 4A). Consistent with our fourth hypothesis, in general, trees sparked by superspreaders were slightly larger than those not sparked by superspreaders (Fig 4A), but there was only a significant difference for COVID-19 trees (P=9.37 · 10^*−*23^, Kruskal-Wallis tests with Bonferroni-Holm corrections). Lack of significance for the other diseases could be due to small sample size. Fig 4B shows partial support for our fifth hypothesis; the proportion of cases in the first generation was significantly positively related to standardized tree size for COVID-19, Ebola, hepatitis A, and Nipah trees based on a linear regression.

**Figure 4:**
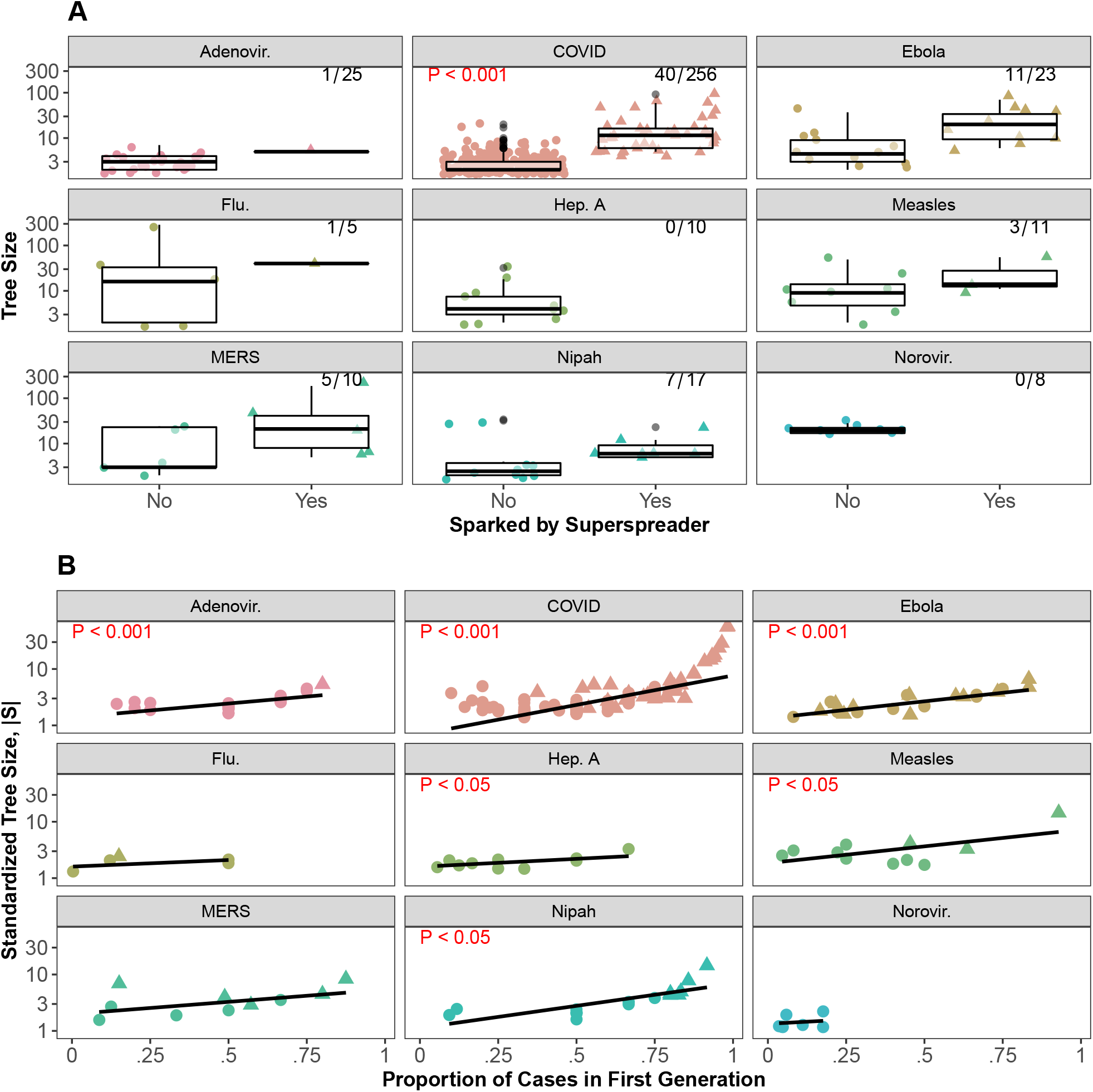
Importance of early spreading events to tree size across diseases. (A) The frequency (top right) and effect of trees sparked by superspreaders on tree size. Red text indicates results of Kruskal-Wallis test with Bonferroni-Holm correction of differences in tree size by superspreader sparking status (only P-values<0.05 shown). (B) Across diseases there is a positive relationship between standardized tree size and proportion of cases in the first generation. Standardized tree size, |*S*|, is calculated by |*S*| = *S*^1*/*(*G−*1)^ where *G* is the number of generations in the tree and *S* is tree size. Lines show linear regression with results in red text (only coefficients with P-values<0.05 shown). Note: y-axis on log10 scale. Trees sparked by superspreaders are represented by triangles, trees not sparked by superspreaders are represented by circles. Points have been jittered for clarity.

Thus, we found limited support for both hypotheses; only COVID-19 trees sparked by superspreaders were significantly larger than trees not sparked by superspreaders and 4/9 diseases had significant positive relationships between proportion of cases in the first generation and standardized tree size. The strong association between proportion of cases in the first generation and standardized tree size for COVID-19 may be a result of higher proportions of cases in the first generation leading to larger tree sizes, but it could also be due to limited detection of onward transmission in COVID-19 trees, decreasing the number of observed generations in a tree. These observations contextualize our finding in the previous analysis, showing that superspreading in COVID-19 trees is occurring early and igniting large outbreaks [18]. The presence of large, superspreader sparked trees with few generations suggests diminished detection of subsequent transmission beyond the first few generations of spread.

### Generation of superspreaders (Hypothesis 6)

Across diseases, there were patterns in the characteristics of individuals that infected superspreaders (Fig 5), which we investigated for Hypothesis 6. More than 50% of superspreaders were infected by other superspreaders in SARS, MERS, influenza, and smallpox trees, whereas COVID-19 superspreaders were infected by another superspreader only about one third of the time. Furthermore, the ratio of observed to expected superspreader-superspreader dyads was greater than one in 13 of 18 trees with two or more superspreaders, indicating that the proportion of superspreaders infected by other superspreaders was greater than expected by chance. Notably, both COVID-19 trees under consideration had large ratios of observed to expected superspreader dyads. Though additional information regarding the contexts in which superspreaders are infected would be required to understand these patterns, these results suggest some non-randomness in generation of superspreaders providing preliminary support for our sixth hypothesis that superspreaders infect other superspreaders.

**Figure 5:**
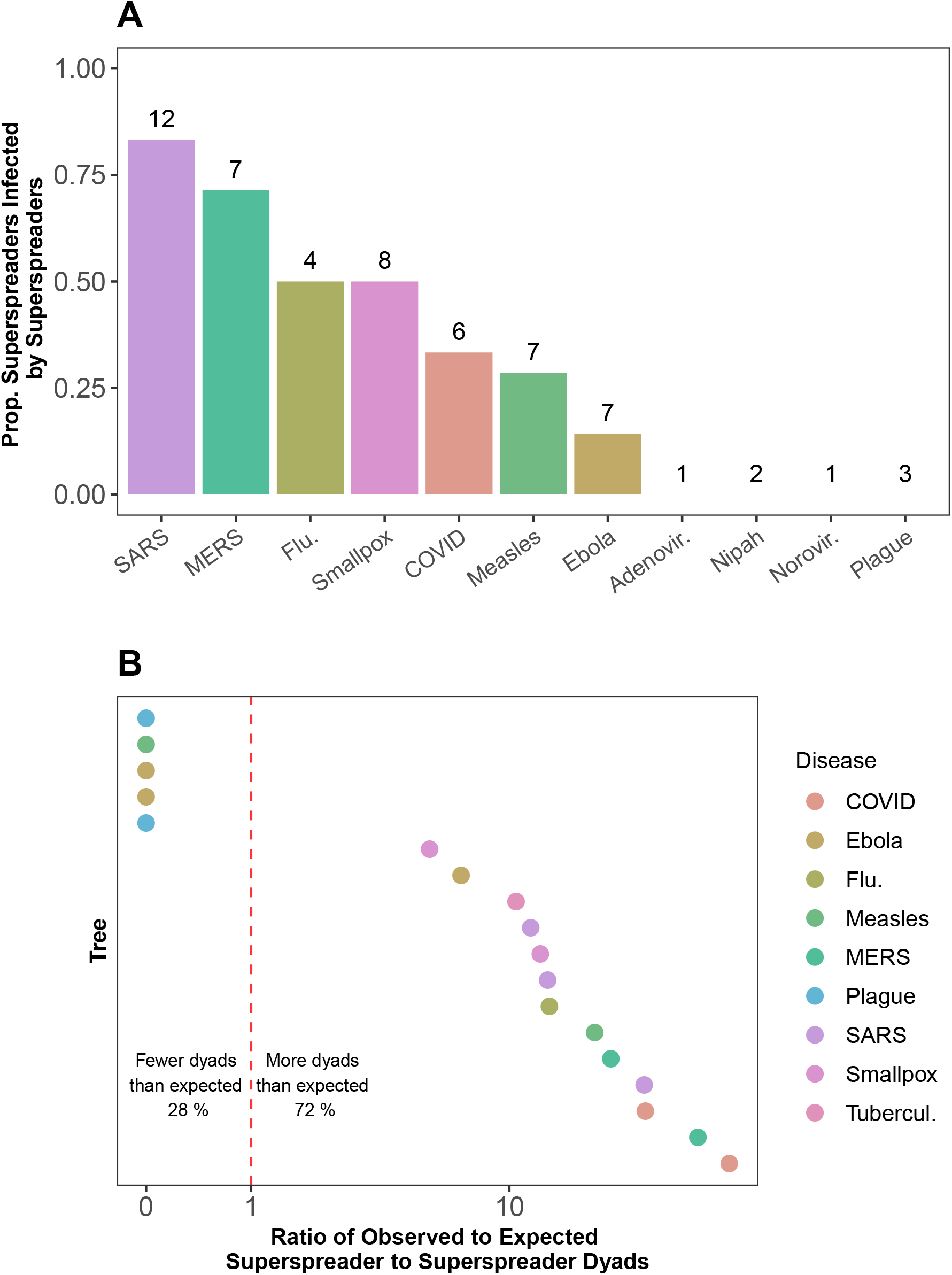
Characteristics of individuals infecting superspreaders. (A) The proportion of superspreaders infected by other superspreaders. Numbers above bars indicate the number of superspreaders for which there was sufficient information about their infector to calculate the proportion. (B) Ratio of observed to expected superspreader-superspreader dyads in trees with more than one superspreader. The expected number of dyads is calculated by *s*(*s −* 1)*/S*, where *s* is the number of superspreaders in the tree and *S* is tree size.

Lastly, we explored the context in which superspreaders were infected when this information was available. As shown in Fig 6, across diseases, superspreaders were most commonly infected at gatherings (e.g., religious, fitness classes) or while traveling. Gatherings and travel allow for contact with many individuals for prolonged duration, potentially facilitating infection with higher viral loads as suggested in [8], although we do not have data to determine if this correlation is a result of biological (higher viral shedding) or behavioral (higher contact rate) differences. Regardless, understanding patterns in the generation of superspreaders is an exciting avenue for future research and our database provides some preliminary data to test new hypotheses.

**Figure 6:**
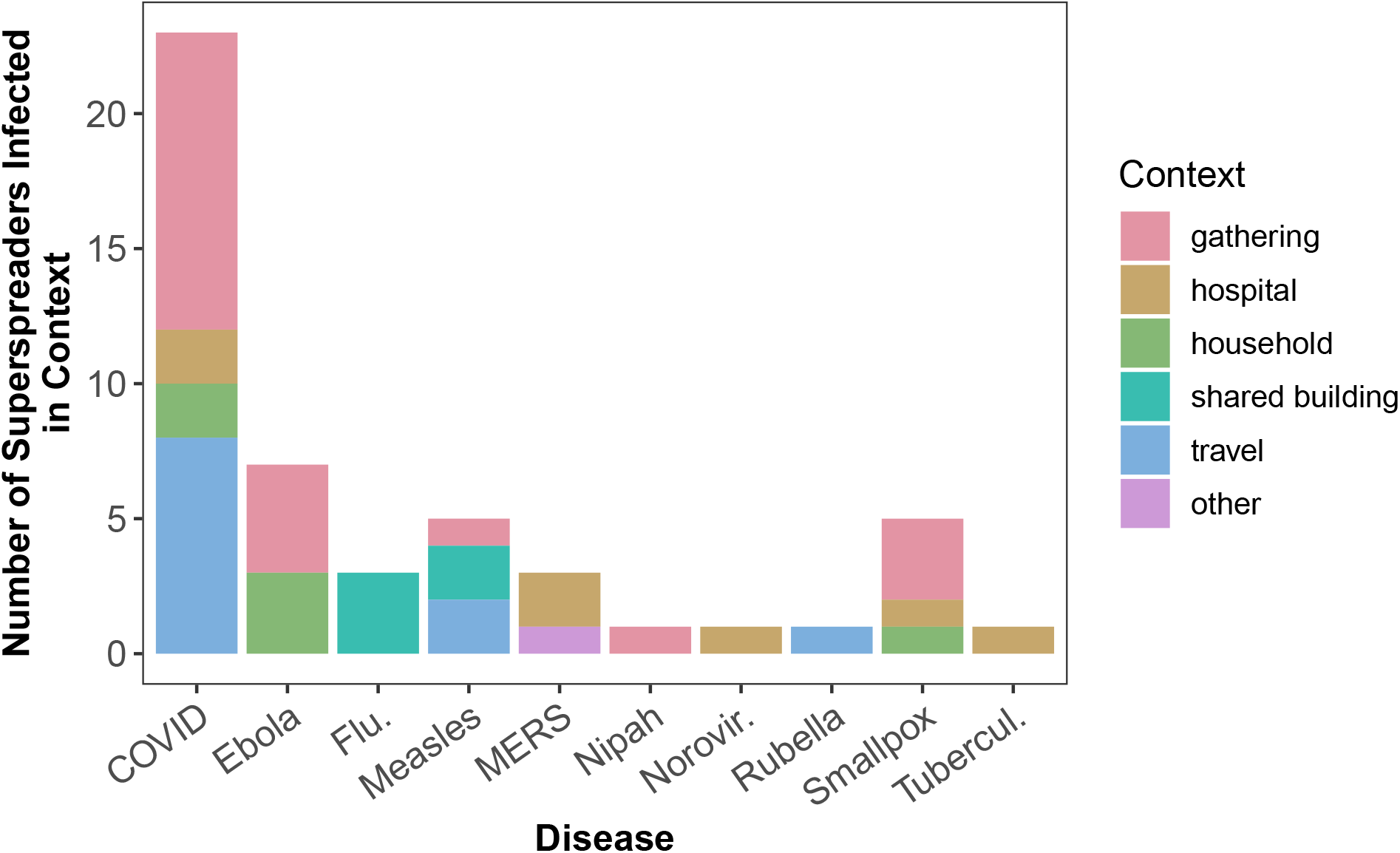
Number of superspreaders infected in various transmission contexts. Shared buildings included worker dormitories and hotels. The category “other” includes a spillover event of MERS to a camel owner.

### Limitations of OutbreakTrees

While `OutbreakTrees` has allowed us to investigate questions about the nature of superspreading, the database has several limitations. First, trees in the database do not constitute a random nor necessarily representative sample of directly-transmitted infectious disease outbreaks. For example, we omitted nearly one hundred reported transmission events and trees due to lack of single infector identification, which undermines the generalization of our findings. Second, although trees are meant to be complete representations of clusters (see inclusion criteria), they are typically a subset from a larger chain of transmission events. For example, Ebola was likely only introduced once in the 2014 outbreak in West Africa, yet we have several separate trees because the transmission events could not all be connected. These limitations should be kept in mind when using the database and analyzing results.

### Intended users & future directions

We envision this database to be used by teachers and scientists. Secondary instruction in mathematics might use `OutbreakTrees` to discuss exponential or geometric growth, concepts outlined in the Common Core State Standards for high school mathematics [19]. College statistics instructors could use the dispersion parameter to discuss the negative binomial distribution in introductory statistics courses. These are simple examples and this list could be easily expanded. With so much current interest in infectious diseases, we believe students would be excited by these topics. In addition to teachers, researchers can use the data in `OutbreakTrees` to study a variety of topics. The data could be used to further explore theory about superspreaders generating new superspreaders, to learn more about how and when outbreaks fade out, or to calculate key statistics to inform public health policy. Trees that have more detail about symptom onset and contact between individuals could be used to test or refine probabilistic reconstruction methods.

`OutbreakTrees` is maintained at OutbreakTrees.ecology.uga.edu where users can visualize and download transmission trees. We will maintain it by entering new trees as they become available and we invite scientists or public health practitioners to submit new trees. In the future, we plan to expand the database to include outbreaks of sexually transmitted infections, transmission trees of animal infectious diseases, and larger trees reconstructed by phylogenetic methods.

## Conclusions

In summary, we developed an open-access database of infectious disease transmission trees, called `OutbreakTrees,` for research and teaching. We illustrated how this database can be used to explore questions surrounding superspreader epidemiology, and we calculated a number of important parameters for COVID-19. In particular, we estimated the dispersion parameter from transmission trees and the value for COVID-19 was in between that of SARS and MERS. Additionally, COVID-19 data supported our hypothesis that transmission trees sparked by superspreaders would be larger than those not sparked by superspreaders. Finally, our analysis provided support for the theory that superspreaders generate other superspreaders, even when controlling for number of secondary infections. The development and release of `OutbreakTrees` highlights the benefits of data sharing and offers a new resource for epidemiologic research.

## Methods

### Data

Transmission trees were collected by searching Google Scholar, Scopus, PubMed, and Google Images for published literature containing graphs of transmission trees or written accounts of transmission events. We used the following terms to find papers containing transmission tree information: “transmission AND (tree OR network OR chain) AND (outbreak OR disease)”, “outbreak investigation”, “contact tracing”, “case report”, and “transmission tree outbreak reconstruction”. We also used the bibliographies of other papers (e.g. [6]) to find more references. With the emergence of COVID-19, we expanded our search for transmission trees to include news articles and pre-prints (e.g. medRxiv.org). For COVID-19, many of the trees were identified with an online database [3]. If trees could not be collected from a public source or if trees did not identify single infectors for each infectee, we contacted the authors of identified documents for further clarification or additional information. We additionally compiled readily available node attributes reported in the tree source. Attributes available for each tree varied but included: age, sex, context of transmission, date of symptom onset, occupation, quarantine status, survival status, location, hospital, ward of hospital or care facility, symptomatic status, duration of exposure to infected individual, whether the edge was probabilistically reconstructed, relationship between individuals, serial interval, immunization status, source of edge (if tree constructed from two sources), and strain or genome sequence. Articles in languages other than English were translated using Google Translate software.

Examples of trees contained in OutbreakTrees are shown in Fig 1.

### Inclusion criteria

For consistency, we required that a tree meet the following criteria for inclusion in the database:

- Trees must have contained two or more individuals; case studies of isolated infected individuals were excluded.
- Trees must represent outbreaks of directly transmitted infectious diseases in humans; trees describing sexually-transmitted, food-borne, vector-borne, or waterborne diseases, as well as diseases in non-humans (e.g. outbreaks among farm animals [20, 21]) were excluded.
- Trees were constructed through epidemiological or probabilistic methods; trees constructed using only genomic or phylogenetic methods were excluded.
- Trees had to report a single infector per infectee (i.e., trees that had multiple possible infectors for any case were excluded). However, if tree topology was unaffected by randomly assigning ambiguous infectors, we included the tree and omitted specific attribute data for the assigned infector.
- Trees were presented as completed investigations; we excluded trees presented as still under ongoing investigation at the time of reporting.

### Data entry

Trees were manually encoded as data.tree [22] objects using relevant information from each source and converted to igraph [23] objects for manipulation and accession. Any assumptions made in entering the tree are listed with the tree in the database (e.g. if an infector is assumed due to nodes obscuring branches, or a case of an ambiguous infector assignment). All scripts to compile trees and analyze data are available at http://github.com/DrakeLab/taube-transmission-trees and tree sources are listed in S1 File.

### Data analysis

We demonstrated how OutbreakTrees can be used to address questions about the role of superspreading in infectious disease transmission through three different analyses.

### Superspreading events across diseases

To evaluate how common superspreading is among different diseases we measured the importance of superspreading across trees and diseases in three ways: by determining for each tree (1) the number of superspreaders based on the definition of Lloyd-Smith et al. [6], (2) the dispersion parameter (*k*), and (3) the cumulative proportion of cases directly and indirectly caused by superspreaders. The dispersion parameter was calculated in R using the `fitdistr` function in the `MASS` package assuming secondary infections followed a negative binomial distribution. There were 9 trees for which we could not calculate a dispersion parameter using this method; all had an offspring distribution of {1, 1, 2} which was not optimizable. Small dispersion parameters indicate more heterogeneous offspring distributions with fewer individuals accounting for the majority of transmission compared with large dispersion parameters. Additionally, we quantified the cumulative proportion of cases downstream from superspreaders as the number of cases which could trace their infection back to a superspreader divided by the total number of secondary cases in the tree (i.e. tree size - 1). This value indicates the contribution of superspreaders as a fraction of all transmission. For this analysis, we used trees consisting of 8 or more cases because cutoffs for superspreading in smaller trees are low (e.g. for a tree with 5 cases, someone who infected 4 cases would be a superspreader) and we sought to compare larger superspreading events. We performed sensitivity analyses for cutoffs of trees with 5 and 10 or more cases (S1 Fig, S2 Fig).

### Tree size and early spreading events

To understand the relationship between early spread and tree size, we analyzed trends in the frequency of trees sparked by superspreaders, tree size, and size of the first generation of cases. To compare the proportion of cases in the first generation with tree size among trees with different numbers of generations, we standardized tree size (*S*) by the number of generations (*G*), not including the index case: |*S*| = *S*^1*/*(*G−*1)^. If the index case was a superspreader [6], then we considered the tree to be sparked by a superspreader. The first generation of cases was defined as those cases infected by the index case. The proportion of cases in the first generation was calculated by dividing the number of cases in the first generation by the total number of cases in the tree. We limited this analysis to diseases with at least five trees.

### Generation of superspreaders

Next, to understand patterns in the individuals who infected superspreaders and the contexts in which superspreaders were infected, we investigated three values. First, we assessed the frequency of superspreaders infected by other superspreaders. We determined the total number of superspreaders infected by another superspreader for each disease, and divided this count by the total number of superspreaders in all trees for that disease. We did not include superspreaders that sparked a tree due to lack of sufficient information about their infector. Second, we calculated the ratio of observed to expected superspreader-superspreader dyads. Superspreader-superspreader dyads occur when one superspreader infects another. Denoting the number of superspreaders by *s* and tree size by *S*, if superspreaders were infected randomly, we would expect *s*(1*− s*)*/S* superspreader-superspreader dyads per tree. Third, we identified the contexts in which superspreaders were infected, when reported, and identified any patterns. Contexts were categorized as either gathering, hospital, household, shared building (worker dormitories or hotel), travel, or other (e.g., spillover from a camel causing MERS infection). If generation of superspreaders is not random but tied to characteristics of the infector and transmission contexts, we would expect to see high frequencies of superspreaders infecting other superspreaders, large ratios of observed to expected superspreader-superspreader dyads, and superspreaders being infected in contexts that encourage individuals to be in close contact with each other.

## Data Availability

All data used in this manuscript are available for download at the following link (https://outbreaktrees.ecology.uga.edu/) and all code used to compile the database is available on GitHub (https://github.com/DrakeLab/taube-transmission-trees).

https://github.com/DrakeLab/taube-transmission-trees

## Acknowledgments

We thank E. Marty for assistance developing the online interface to OutbreakTrees.

