## Supplementary figures and images for "An open-access database of infectious disease transmission trees to explore superspreader epidemiology"

### S1Fig

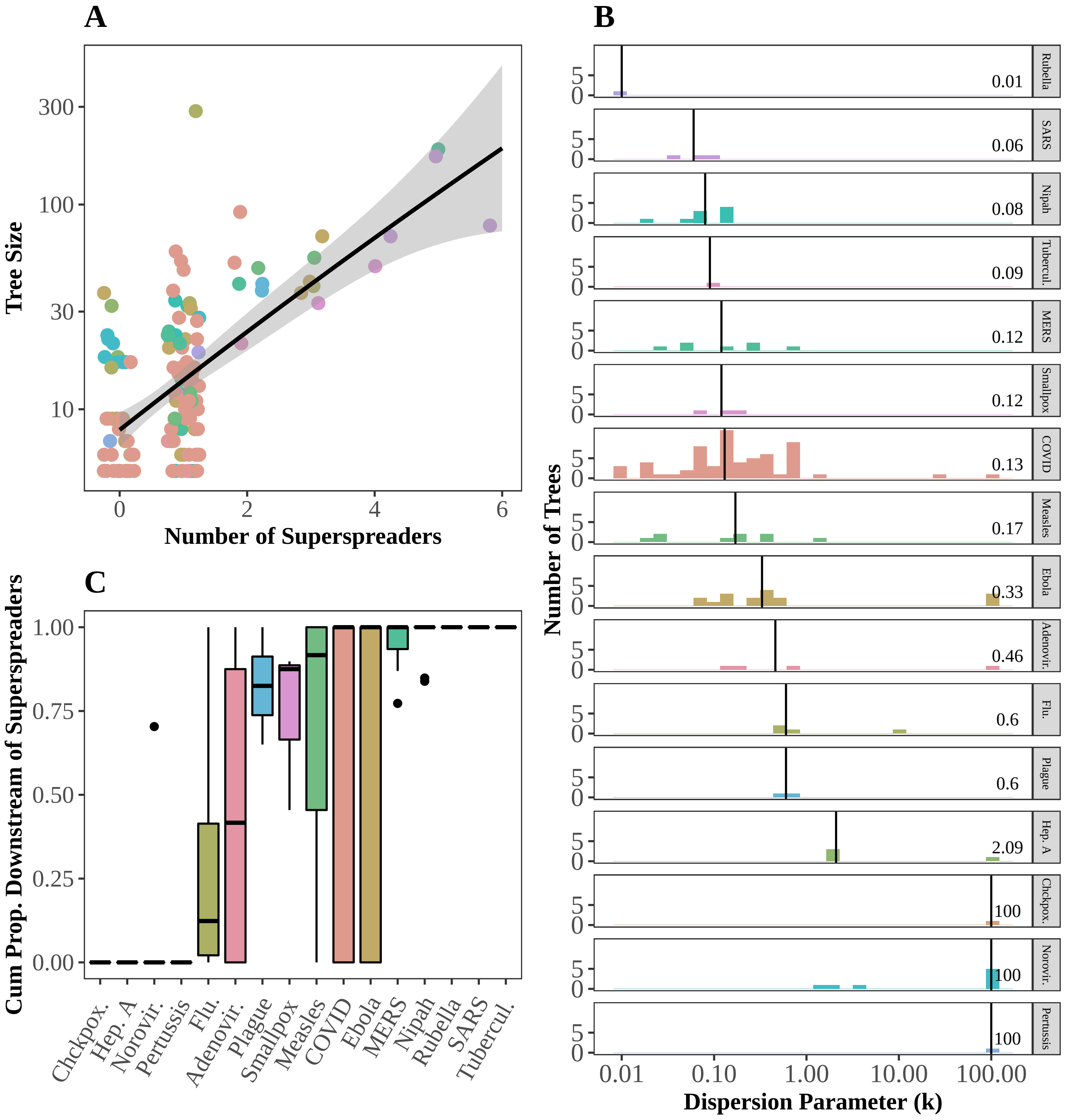

### S2Fig

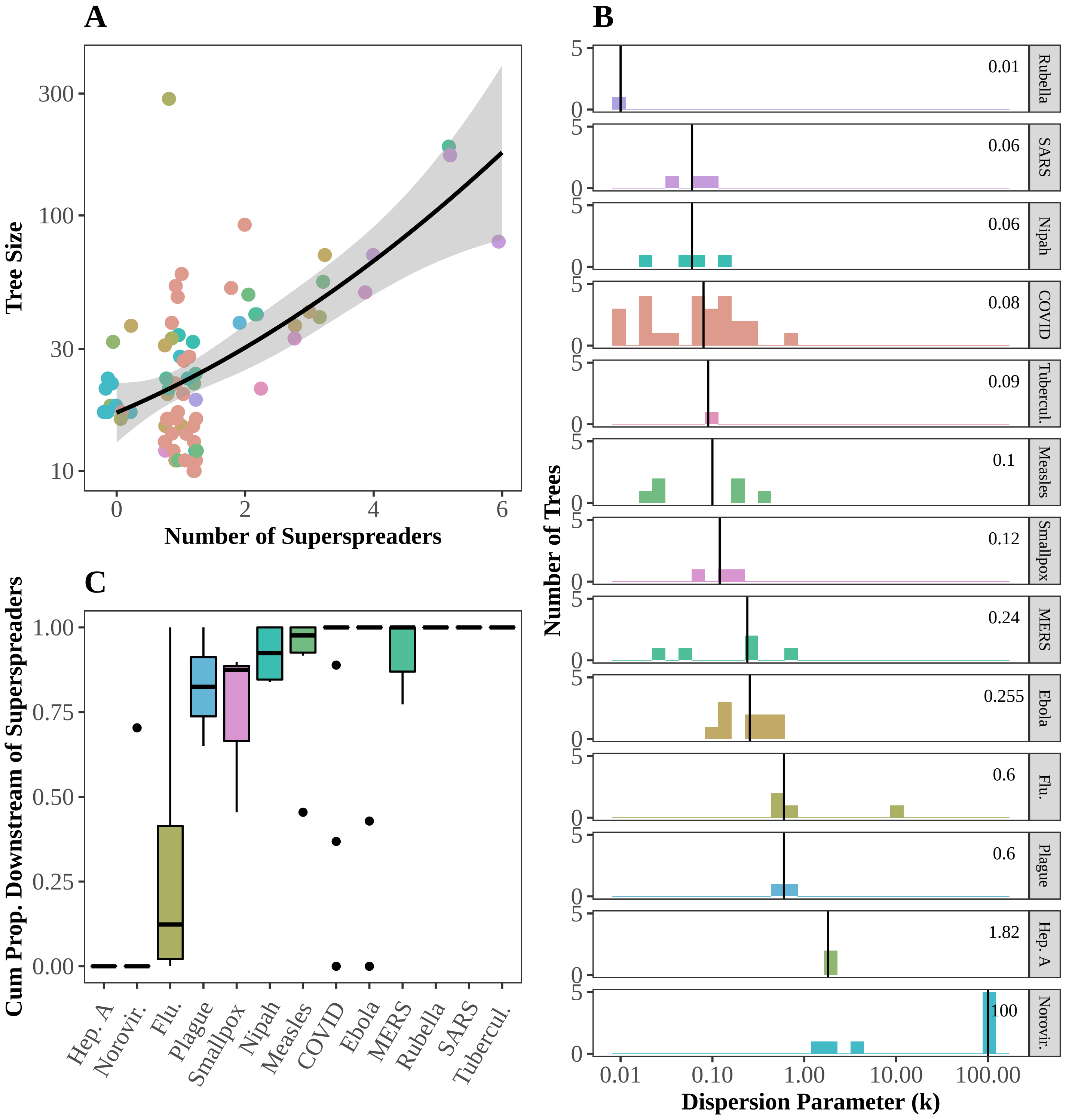

### S3Fig

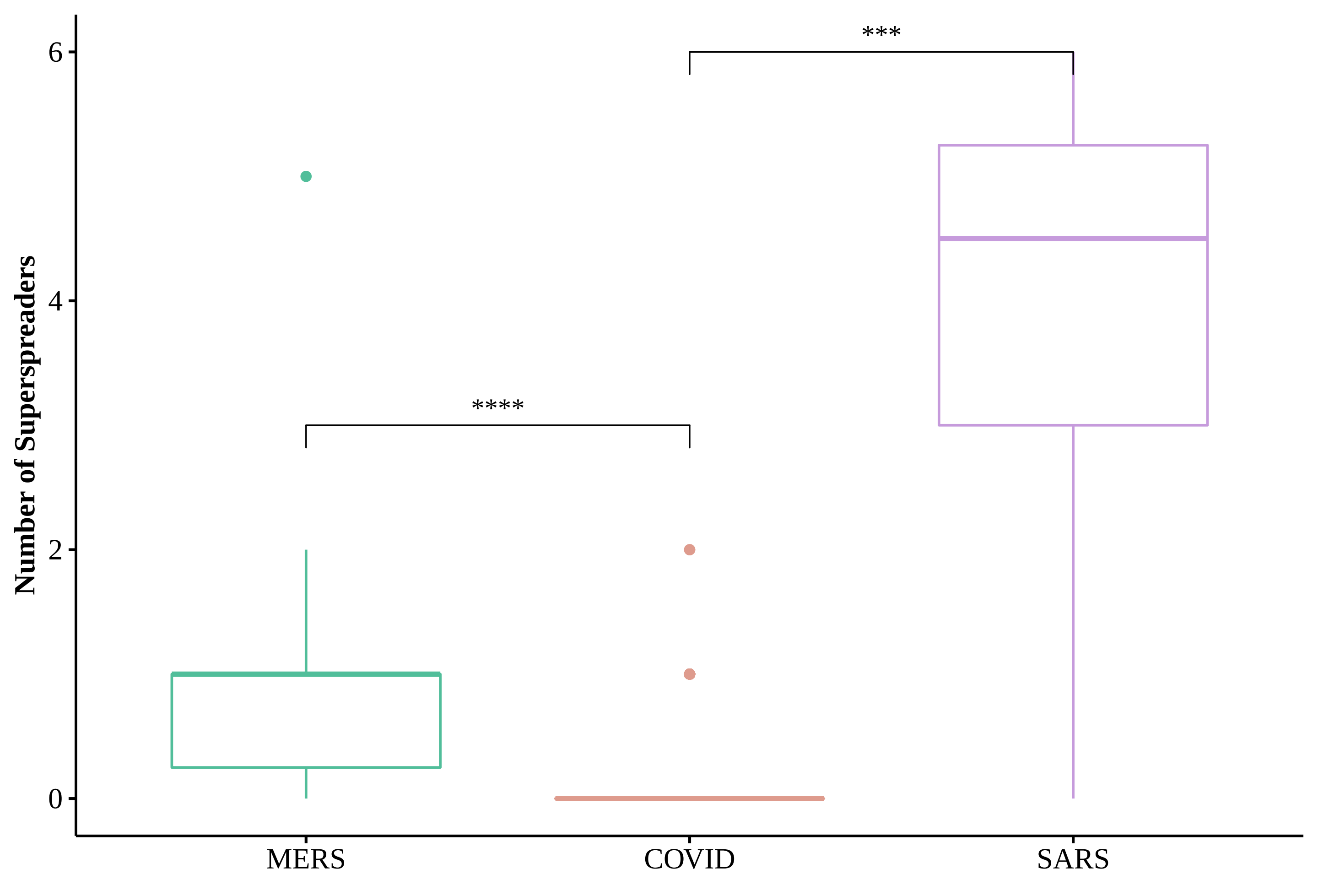
