## Supplementary material for "An open-access database of infectious disease transmission trees to explore superspreader epidemiology": S1File

http://country.cnr.cn/gundong/20200205/t20200205_524961561.shtml

http://hlj.people.com.cn/GB/n2/2020/0205/c220024-33767665.html

http://ncirs.org.au/sites/default/files/2020-04/NCIRS%20NSW%20Schools%20COVID_Summary_FINAL%20public_26%20April%202020.pdf

http://news.china.com.cn/2020-01/30/content_75657770.htm

http://web.archive.org/web/20200420071139/https://edition.cnn.com/2020/03/24/europe/austria-ski-resort-ischgl-coronavirus-intl/

http://web.archive.org/web/20200420071353/https://www.aljazeera.com/news/2020/03/infected-16-berlin-nightclub-coronavirus-fears-grow-200310132859234.html

http://web.archive.org/web/20200420071548/https://www.theguardian.com/world/2020/feb/26/coronavirus-inquiry-opens-into-hospitals-at-centre-of-italy-outbreak

http://web.archive.org/web/20200420072800/https://wsbt.com/news/local/first-cases-of-coronavirus-confirmed-in-berrien-county

http://www.koreabiomed.com/news/articleView.html?idxno=8291

https://btvnovinite.bg/bulgaria/izsledvat-oshte-30-dushi-kontaktuvali-sas-zarazenite-s-koronavirus-na-tarzhestvo-v-lom.html

https://coronavirus.ohio.gov/wps/portal/gov/covid-19/resources/news-releases-news-you-can-use/covid-19-update-08-04-20

https://dnes.dir.bg/obshtestvo/otvaryat-shivashkata-fabrika-ognishte-na-covid-19-v-pleven

https://dph.georgia.gov/press-releases/2020-03-02/gov-kemp-officials-confirm-two-cases-covid-19-georgia

https://en.vietnamplus.vn/vietnam-reports-two-more-coronavirus-infection-cases/168264.vnp

https://vietnaminsider.vn/vietnam-reports-a-new-case-of-coronavirus-infection-in-nha-trang/

https://www.cbc.ca/news/canada/british-columbia/covid-19-coronavirus-update-1.5572681

https://www.cdc.gov.tw/Bulletin/Detail/1T3s2mxcCjqlE0MczXRD8w?typeid=9

https://www.info.gov.hk/gia/general/202002/02/P2020020200627.htm

https://www.wsj.com/articles/how-one-singapore-sales-conference-spread-coronavirus-around-the-world-11582299129

Hu, Z., Song, C., Xu, C., Jin, G., Chen, Y., Xu, X., ... & Wang, J. (2020). Clinical characteristics of 24 asymptomatic infections with COVID-19 screened among close contacts in Nanjing, C hina. *Science China Life Sciences*, 63(5), 706-711.

Huang, C., Wang, Y., Li, X., Ren, L., Zhao, J., Hu, Y., ... & Cheng, Z. (2020). Clinical features of patients infected with 2019 novel coronavirus in Wuhan, China. *The lancet*, 395(10223), 497-506.

Hunter, J. C., Nguyen, D., Aden, B., Al Bandar, Z., Al Dhaheri, W., Elkheir, K. A., ... & Al Kaabi, N. (2016). Transmission of Middle East respiratory syndrome coronavirus infections in healthcare settings, Abu Dhabi. *Emerging infectious diseases*, 22(4), 647.

Infectious Diseases Team and Director, South Eastern Sydney Public Health Unit. (2001). Outbreak report: measles cluster in south-eastern Sydney with transmission in a general practice waiting room. *Communicable diseases intelligence quarterly report*, 25(1).

Jang, S., Han, S. H., & Rhee, J. Y. (2020). Cluster of coronavirus disease associated with fitness dance classes, South Korea. *Emerging infectious diseases*, 26(8).

Ji, L. N., Chao, S., Wang, Y. J., Li, X. J., Mu, X. D., Lin, M. G., & Jiang, R. M. (2020). Clinical features of pediatric patients with COVID-19: a report of two family cluster cases. *World Journal of Pediatrics*, 1-4.

Jing, Q. L., Liu, M. J., Zhang, Z. B., Fang, L. Q., Yuan, J., Zhang, A. R., ... & Kenah, E. (2020). Household secondary attack rate of COVID-19 and associated determinants in Guangzhou, China: a retrospective cohort study. *The Lancet Infectious Diseases*.

Kam, K. Q., Yung, C. F., Cui, L., Tzer Pin Lin, R., Mak, T. M., Maiwald, M., ... & Thoon, K. C. (2020). A well infant with coronavirus disease 2019 with high viral load. *Clinical Infectious Diseases*.

Khan, M. H. R. (2020). COVID-19-19 (Coronavirus) in Bangladesh (No. 63-08062020). Report.

Ki, M. (2020). Epidemiologic characteristics of early cases with 2019 novel coronavirus (2019-nCoV) disease in Korea. *Epidemiology and health*, 42.

Komabayashi, K., Seto, J., Tanaka, S., Suzuki, Y., Ikeda, T., Onuki, N., ... & Mizuta, K. (2018). The largest measles outbreak, including 38 modified measles and 22 typical measles cases, Yamagata, Japan, 2017 in its elimination era. *Japanese Journal of Infectious Diseases*, JJID-2018.

L. T., Nguyen, T. V., Luong, Q. C., Nguyen, T. V., Nguyen, H. T., Le, H. Q., ... & Pham, Q. D. (2020). Importation and human-to-human transmission of a novel coronavirus in Vietnam. *New England Journal of Medicine*, 382(9), 872-874.

Lai, C. C., Wang, C. Y., Wang, Y. H., Hsueh, S. C., Ko, W. C., & Hsueh, P. R. (2020). Global epidemiology of coronavirus disease 2019: disease incidence, daily cumulative index, mortality, and their association with country healthcare resources and economic status. *International journal of antimicrobial agents*, 105946.

Le, H. T., Nguyen, L. V., Tran, D. M., Do, H. T., Tran, H. T., Le, Y. T., & Phan, P. H. (2020). The first infant case of COVID-19 acquired from a secondary transmission in Vietnam. *The Lancet Child & Adolescent Health*, 4(5), 405-406.

Li, G. G., Lv, Z., Wang, Y. S., Li, J. F., Feng, L. F., Wang, M. F., ... & Pan, X. L. (2020). Retrospective

Analysis of 2019-nCov-Infected Cases in Dongyang, Southeastern China. *Canadian Journal of Infectious Diseases and Medical Microbiology*, 2020.

Li, P., Fu, J. B., Li, K. F., Chen, Y., Wang, H. L., Liu, L. J., ... & Tong, Z. D. (2020). Transmission of COVID-19 in the terminal stage of incubation period: a familial cluster. *International Journal of Infectious Diseases.*

Li, Q., Guan, X., Wu, P., Wang, X., Zhou, L., Tong, Y., ... & Xing, X. (2020). Early transmission dynamics in Wuhan, China, of novel coronavirus–infected pneumonia. *New England Journal of Medicine.*

Lim, J., Jeon, S., Shin, H. Y., Kim, M. J., Seong, Y. M., Lee, W. J., ... & Park, S. J. (2020). Case of the index patient who caused tertiary transmission of coronavirus disease 2019 in Korea: The application of lopinavir/ritonavir for the treatment of COVID-19 pneumonia monitored by quantitative RT-PCR. *Journal of Korean Medical Science*, 35(6).

Liu, M., Liu, L., Li, P., Ding, Y., Wu, T., Tang, W., ... & Cao, G. (2020). Presymptomatic transmission and diverse progression of familial clustering COVID-19 cases in Zhoushan, China.

Liu, Y. C., Liao, C. H., Chang, C. F., Chou, C. C., & Lin, Y. R. (2020). A locally transmitted case of SARS-CoV-2 infection in Taiwan. *New England Journal of Medicine*, 382(11), 1070-1072.

Liu, Y. F., Li, J. M., Zhou, P. H., Liu, J., Dong, X. C., Lyu, J., & Zhang, Y. (2020). Analysis on cluster cases of COVID-19 in Tianjin. *Zhonghua Liu Xing Bing Xue Za Zhi= Zhonghua Liuxingbingxue Zazhi*, 41(5), 654-657.

Luo, C., Ma, Y., Jiang, P., Zhang, T., & Yin, F. (2020). The construction and visualization of the transmission networks for COVID-19: A potential solution for contact tracing and assessments of epidemics.

Ma, X., Su, L., Zhang, Y., Zhang, X., Gai, Z., & Zhang, Z. (2020). Do children need a longer time to shed SARS-CoV-2 in stool than adults?. *Journal of Microbiology, Immunology and Infection*.

Mailles, A., Blanckaert, K., Chaud, P., Van der Werf, S., Lina, B., Caro, V., ... & Paty, M. C. (2013). First cases of Middle East Respiratory Syndrome Coronavirus (MERS-CoV) infections in France, investigations and implications for the prevention of human-to-human transmission, France, May 2013. *Eurosurveillance*, 18(24), 20502.

Nikolay, B., Salje, H., Hossain, M. J., Khan, A. D., Sazzad, H. M., Rahman, M., ... & Nichol, S. T. (2019). Transmission of Nipah Virus—14 Years of Investigations in Bangladesh. *New England Journal of Medicine,* 380(19), 1804-1814.

Nishiura, H., Mizumoto, K., & Asai, Y. (2017). Assessing the transmission dynamics of measles in Japan, 2016. *Epidemics*, 20, 67-72.

Nishiura, H., Schwehm, M., Kakehashi, M., & Eichner, M. (2006). Transmission potential of primary pneumonic plague: time inhomogeneous evaluation based on historical documents of the transmission network. *Journal of Epidemiology & Community Health*, 60(7), 640-645.

Nissen, K., Hagbom, M., Krambrich, J., Akaberi, D., Sharma, S., Ling, J., ... & Salaneck, E. (2020). Presymptomatic viral shedding and infective ability of Severe Acute Respiratory Syndrome coronavirus 2.

Nkoghe, D., Kone, M. L., Yada, A., & Leroy, E. (2011). A limited outbreak of Ebola haemorrhagic fever in Etoumbi, Republic of Congo, 2005. *Transactions of the Royal Society of Tropical Medicine and Hygiene*, 105(8), 466-472.

Pallivalappil, B., Ali, A., Thulaseedharan, N. K., Karadan, U., Chellenton, J., Dipu, K. P., ... & Kumar, G. S. (2020). Dissecting an outbreak: A clinico-epidemiological study of Nipah virus infection in Kerala, India, 2018. *Journal of Global Infectious Diseases*, 12(1), 21.

Park, J. Y., Han, M. S., Park, K. U., Kim, J. Y., & Choi, E. H. (2020). First pediatric case of coronavirus disease 2019 in Korea. Journal of Korean medical science, 35(11).

Qi, X., Qian, Y. H., Bao, C. J., Guo, X. L., Cui, L. B., Tang, F. Y., ... & Xu, K. (2013). Probable person to person transmission of novel avian influenza A (H7N9) virus in Eastern China, 2013: epidemiological investigation. *BMJ*, 347, f4752.

Qiu, C., Deng, Z., Xiao, Q., Shu, Y., Deng, Y., Wang, H., ... & Zhou, J. (2020). Transmission and clinical characteristics of coronavirus disease 2019 in 104 outside-Wuhan patients, China. *Journal of Medical Virology.*

Rickman, H. M., Rampling, T., Shaw, K., Martinez-Garcia, G., Hail, L., Coen, P., ... & Houlihan, C. F. (2020). Nosocomial transmission of COVID-19: a retrospective study of 66 hospital-acquired cases in a London teaching hospital. *Clinical Infectious Diseases*.

Sanchez-Codez, M., Hunt, W. G., Watson, J., & Mejias, A. (2020). Hepatitis in children with tuberculosis: a case report and review of the literature. *BMC Pulmonary Medicine*, 20(1), 1-4.

Sansone, M. (2020). Epidemiology of viral respiratory infections with focus on in-hospital influenza transmission.

Shen, Z., Ning, F., Zhou, W., He, X., Lin, C., Chin, D. P., ... & Schuchat, A. (2004). Superspreading sars events, Beijing, 2003. *Emerging infectious diseases*, 10(2), 256.

Shimizu, K., Kinoshita, R., Yoshii, K., Akhmetzhanov, A. R., Jung, S., Lee, H., & Nishiura, H. (2018). An investigation of a measles outbreak in Japan and China, Taiwan, China, March–May 2018. Western Pacific surveillance and response journal: *WPSAR*, 9(3), 25.

Simpson, E. H., & Breuer, J. (2002). Studies on shingles, is the virus ordinary chickenpox virus?. *Reviews in Medical Virology*, 12(1), 5.

Singh, R., & Singh, P. K. (2020). Connecting the Dots of COVID-19 Transmissions in India. *arXiv preprint* arXiv:2004.07610.

Small, P. M., Hopewell, P. C., Singh, S. P., Paz, A., Parsonnet, J., Ruston, D. C., ... & Schoolnik, G. K. (1994). The epidemiology of tuberculosis in San Francisco--a population-based study using conventional and molecular methods. *New England Journal of Medicine*, 330(24), 1703-1709.

Soetens, L., Klinkenberg, D., Swaan, C., Hahné, S., & Wallinga, J. (2018). Real-time Estimation of Epidemiologic Parameters from Contact Tracing Data During an Emerging Infectious Disease Outbreak. *Epidemiology*, 29(2), 230-236.

Su, L., Ma, X., Yu, H., Zhang, Z., Bian, P., Han, Y., ... & Zhang, Z. (2020). The different clinical characteristics of corona virus disease cases between children and their families in China–the character of children with COVID-19. *Emerging microbes & infections*, 9(1), 707-713.

Sukhrie, F. H., Teunis, P., Vennema, H., Copra, C., Thijs Beersma, M. F., Bogerman, J., & Koopmans, M. (2012). Nosocomial transmission of norovirus is mainly caused by symptomatic cases. *Clinical Infectious Diseases*, 54(7), 931-937.

Teunis, P., Heijne, J. C., Sukhrie, F., van Eijkeren, J., Koopmans, M., & Kretzschmar, M. (2013). Infectious disease transmission as a forensic problem: who infected whom?. *Journal of the Royal Society Interface,* 10(81), 20120955.

Thanh, H. N., Van, T. N., Thu, H. N. T., Van, B. N., Thanh, B. D., Thu, H. P. T., ... & Nguyen, T. A. (2020). Outbreak investigation for COVID-19 in northern Vietnam. *The Lancet Infectious Diseases,* 20(5), 535-536.

Tieh, T. H., Landauer, E., Miyagawa, F., Kobayashi, G., & Okayasu, G. (1948). Primary pneumonic plague in Mukden, 1946, and report of 39 cases with 3 recoveries. *The Journal of infectious diseases*, 82(1), 52-58.

Tong, Z. D., Tang, A., Li, K. F., Li, P., Wang, H. L., Yi, J. P., ... & Yan, J. B. (2020). Potential presymptomatic transmission of SARS-CoV-2, Zhejiang province, China, 2020*. Emerging infectious diseases*, 26(5), 1052.

Uyaroğlu, O. A., Başaran, N. Ç., Özışık, L., Taş, Z., Dizman, G. T., İnkaya, A. Ç., ... & Güven, G. S. (2020). First Confirmed Cases of 2019 Novel Coronavirus in a University Hospital in Turkey: Housemate Internists. *Acta Medica*.

Valencia, C., Bah, H., Fatoumata, B., Rodier, G., Diallo, B., Koné, M., ... & Jansa, J. (2017). Network visualization for outbreak response: Mapping the Ebola Virus Disease (EVD) chains of transmission in N'Zérékoré, Guinea. *Journal of Infection*, 74(3), 294-301.

Wang, S. X., Li, Y. M., Sun, B. C., Zhang, S. W., Zhao, W. H., Wei, M. T., ... & Cheung, A. C. (2006). The SARS outbreak in a general hospital in Tianjin, China–the case of super-spreader. *Epidemiology & Infection*, 134(4), 786-791.

Weinstein, I. (1947). An outbreak of smallpox in New York City. *American Journal of Public Health and the Nations Health*, 37(11), 1376-1384.

Wong, C. H., Lam, W. H., Lam, H. Y., Lam, T. S., Ho, L. M. R., Leung, Y. H., ... & Chuang, S. K. (2020). Investigation and control of measles outbreak in the Hong Kong International Airport, 2019. *Western Pacific Surveillance and Response*, 11(2).

World Health Organization. (2019). Middle East respiratory syndrome coronavirus (MERS-CoV) - The Kingdom of Saudi Arabia. Retrieved August 6, 2019 from: https://www.who.int/csr/don/24-April-2019-mers-saudi-arabia/en/

Zhang, J., Tian, S., Lou, J., & Chen, Y. (2020). Familial cluster of COVID-19 infection from an asymptomatic. *Critical Care*, 24, 1-3.

Zhang, X. S., & De Angelis, D. (2015). Construction of the influenza A virus transmission tree in a college-based population: co-transmission and interactions between influenza A viruses. BMC infectious diseases, 16(1), 38. Liu, W., Li, Z. D., Tang, F., Wei, M. T., Tong, Y. G., Zhang, L., ... & Zhan, L. (2010). Mixed infections of pandemic H1N1 and seasonal H3N2 viruses in 1 outbreak. *Clinical Infectious Diseases*, 50(10), 1359-1365.

Zhang, X. S., & Iacono, G. L. (2018). Estimating human-to-human transmissibility of hepatitis A virus in an outbreak at an elementary school in China, 2011. *PloS one*, 13(9), e0204201.

Zhang, X. S., Pebody, R., Charlett, A., de Angelis, D., Birrell, P., Kang, H., ... & Choi, Y. H. (2017). Estimating and modelling the transmissibility of Middle East Respiratory Syndrome CoronaVirus during the 2015 outbreak in the Republic of Korea. *Influenza and other respiratory viruses*, 11(5), 434-444.

Zhang, X. S., Smith, A., Patel, B., Anderson, C., Pomeroy, L., Higgins, G., ... & Atchison, C. (2020).

New approaches to controlling an outbreak of chickenpox in a large immigration detention setting in England: the role of serological testing and mathematical modelling. *Epidemiology & Infection*, 148.

Zhao, H., Li, B. S., Xia, Y., Zhou, H. L., Li, T. R., Zeng, Y., ... & Li, Q. (2020). Investigation of transmission chain of a cluster COVID-19 cases. *Zhonghua liu xing bing xue za zhi= Zhonghua liuxingbingxue zazhi*, 41, E064.
